# Evaluating tramadol utilization and patterns by county-level social determinants of health characteristics from 2015 to 2022

**DOI:** 10.1101/2025.11.03.25339433

**Authors:** Chisom Eze, Marie Abate, Gordon Smith, Zheng Dai, Nathan Wood, Mohammad Al-Mamun

**Affiliations:** Department of Pharmaceutical Systems and Policy, School of Pharmacy, West Virginia University, Morgantown, West Virginia, USA; Department of Clinical Pharmacy, School of Pharmacy, West Virginia University, Morgantown, West Virginia, USA; Department of Epidemiology and Biostatistics, School of Public Health, West Virginia University, Morgantown, West Virginia, USA; Health Affairs Institute, Health Sciences Center, West Virginia University, Morgantown, West Virginia, USA; West Virginia Board of Pharmacy, Charleston, West Virginia, USA

## Abstract

**Background:** Tramadol, an opioid analgesic, is liable to abuse and implicated in drug overdose deaths in the U.S. However, tramadol use and its driving factors have not been intricately examined. This study aims to evaluate the tramadol utilization trends by dosage and county, and social determinants of health (SDoH)-related factors associated with the utilization rates.

**Methods:** The retrospective study utilized 2015-2022 WV controlled substances monitoring program, CDC opioid dispensing rate, and County Health Ranking and Roadmaps data. Average tramadol daily dose and morphine milligram equivalent were calculated. Annual tramadol dispensing and use rate per 100 population were calculated for WV and each county. Pooled Poisson regression model was used to analyze the relationship between tramadol dispensing rate, opioid dispensing rate, and SDoH variables.

**Results:** Tramadol dispensing rate declined by 35% (2015-2022), but varied within counties with Grant (34.10), McDowell (28.91), and Wyoming (26.67) average annual rates exceeding the overall WV rate (17.90). High tramadol dispensing rate was associated with a high percentage of the population with poor/fair health (**β=0.07, p=0.01)**, physically inactive (**β=0.10, p=0.0003**), uninsured (**β=0.09, p=0.001)**, and elevated primary care provider (PCP) rate (**β=0.10, p=0.0003**) and opioid dispensing rate (**β=0.14, p<0.0001**).

**Conclusion:** Our study found heterogenous trends of tramadol dispensing rate within WV and was associated with county-wise health status, physical inactivity, insurance, PCP, and opioid dispensing rates. Considering these factors in local surveillance might improve health, and reduce disease burden, drug, and health resource utilization.

## 1.1 Introduction

Tramadol is an opioid analgesic for managing moderate to severe pain (1). It is considered relatively safer than other opioid analgesics due to a lower addiction potential and adverse effects (2). From 2007 to 2008, U.S. tramadol annual prescriptions exceeded those of other opioids, excluding hydrocodone (3).

Emergency department (ED) visits for non-medical use of tramadol and reports of its toxic exposures increased over three-fold from 2004 to 2010/2011 (3). In 2014, tramadol was placed in Schedule IV of the Controlled Substances Act (CSA) (1,4,5).

Despite tramadol’s scheduling, its use, misuse, and related harm appear to be increasing. In the U.S., between 2007 and 2018, the number of ED visits with tramadol prescription increased by about 3.08 million (67% increase), while overall opioid use decreased from 28.4% of ED visits in 2007 to 17.9% in 2018 (6). In 2022, 28.7 million prescriptions for tramadol-containing products were written across the U.S (7). In 2022, the National Survey of Drug Use and Health (NSUDH) reported that 1.4 million persons out of 8.5 million who misused a prescription pain medicine misused tramadol and 41.3% of misused pain medications were obtained through prescriptions or stolen from a healthcare provider (8). Moreover, the National Forensic Laboratory Information System drug annual reports recorded elevated tramadol-involved fatalities from 1,093 to 14,856 cases between 2008 and 2021 (9,10).

In West Virginia (WV), prevalent tramadol use among the commercially insured increased from 2014 to 2021 (11). Since 2016, WV Medicaid placed an age limit on tramadol-containing agents and required prior authorization from Medicaid Rational Drug Therapy Program (12). It is worth to mention that tramadol utilization in WV is not well explored, although it has high chronic disease burden and adults with poor health (13–15). Moreover, the prevalence of these conditions varies widely among counties (13,16,17) and coupled with socio-determinants of health, might result in variable tramadol utilization across counties.

Tramadol utilization demonstrating demographic and regional differences in drug use remains unexamined. Thus, the objective was to evaluate WV county-wise trends in tramadol utilization and social determinant of health (SDoH) factors related to high or low usage post-controlled substance scheduling. An inverse relationship between tramadol and overall opioid dispensing rates is hypothesized. Further, we hypothesized an association between county-wise tramadol use variations and various SDoH factors. Our findings are crucial for understanding tramadol utilization changes since its schedule IV designation by identifying counties with higher utilization rates for closer monitoring, establishing associated predictors for high utilization, and highlighting regions of interest for efficient resource disbursement.

## 2.1 Methods

### 2.1.1 Study Population

The retrospective study sourced data from the 2015 - 2022 WV Controlled Substance Monitoring Program (WVCSMP) data. The WVCSMP is designed for collecting data on all controlled substances dispensed in WV (18). The data was assessed for this study in June 2024. Exemption was obtained from the West Virginia University Institutional Review Board (IRB) due to the deidentified nature of the data. The authors had no access to information that could identify individual participants at any point in the study. Non-institutionalized WV patients who filled ≥1 tramadol prescription each year were included. The WVCSMP data contains de-identified patient demographic, prescriber, and pharmacy-specific information. Drug-specific information includes each drug’s National Drug Code (NDC), name, strength, days supplied, and schedule. Annual 2015 to 2022 opioid dispensing rates (ODR) for WV and its 55 counties were obtained from the Centers for Disease Control (CDC) ODR maps (19). Yearly population estimates for WV and each county were obtained from the Census Bureau website (20). County SDoH variables were extracted from the University of Washington, County Health Rankings and Roadmap (21).

### 2.1.2 Measures

An average daily dose of tramadol was calculated per patient from the total dose and days supplied. The average daily Morphine Milligram Equivalent (MME) of tramadol was derived yearly using the formula:

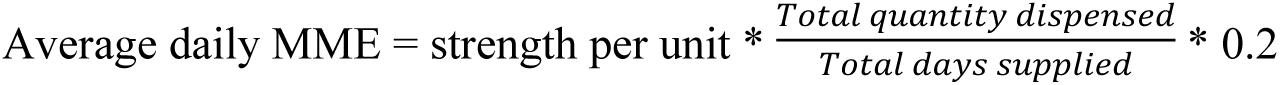 (Tramadol conversion factor)^16^

The tramadol conversion factor, 0.2, was used across the years to keep the MME calculation uniform. Drug dose and MME categories were created based on daily intake of ≤400mg or >400mg which is the maximum recommended dose of tramadol (1), and ≤50 MME or >50 MME of tramadol, respectively, since patients do not generally gain additional benefit from receiving >50 MME of an opioid daily (22). Yearly WV and county-wise tramadol dispensing rates (TDR) per 100 persons were calculated from the annual total number of tramadol prescriptions and the respective population. Annual WV and county-wise tramadol use per 100 persons were also calculated using the total number of distinct patients filling a tramadol prescription in a year and the respective population. Percentages of tramadol prescriptions written by an out-of-state prescriber and tramadol prescriptions filled in another state were also determined. County-wise median annual TDR and ODR were determined. Based on the median value for each variable, counties were categorized as high (>median) or low (≤median) TDR and ODR. Tramadol monthly co-prescription with other prescription medicines including other opioids, benzodiazepines, sedatives, barbiturates, gabapentinoids, and stimulants were derived by number and types of medications. Other variables from the WVCSMP data included age group, gender, and insurance type.

County SDoH variables included proportions of county residents with poor or fair health and physical inactivity, primary care provider (PCP), income ratio, percentage of excessive drinking, smokers, unemployed, uninsured, completed high school, with some college education, severe housing problem, Asians, Hispanics, non-Hispanic Whites, Black/African Americans, American Indian/Native Americans, females, <18 years, and ≥65 years. SDoH variables were selected based on prior literature. Based on the distribution of each SDoH variable, the mean or median was determined. The counties were categorized as having a value greater than or less than/equal to the mean/median value for each variable. However, before categorizing the variables, missing values in the percentage of excessive drinking and PCP rates were imputed by predictive mean matching using the *MICE* package in R Programming software (23).

### 2.1.3 Statistical Analyses

Counts and percentages were obtained for patient characteristics by tramadol dose and MME categories. A Chi-square test was used for group comparisons within each category. Graphical comparison of TDR and ODR across the years were also conducted. To understand county-wise variations in prescription patterns, maps were generated to illustrate and compare county average TDR with the respective average ODR. Tramadol co-prescription by number and type of other medications were represented graphically.

A pooled Poisson regression model with Lasso Regularization was utilized to model the relationship between yearly county TDR as the dependent variable and SDoH factors and ODR as the independent variables. The level of significance (p-value) was set at 0.05. A region-based analysis was conducted by dividing the counties into either northern or southern WV regions (24), to determine the broader effect of geography on the outcome estimates. The statistical models’ appropriateness and fit were tested using an overdispersion test and residual diagnostics. A non-significant p-value for the overdispersion test and residual plot with no obvious patterns confirmed the models’ appropriateness. Data cleaning and analysis were done using R programming version 4.4.0 (25).

## 3.1 Results

### 3.1.1 Tramadol utilization by dosage and MMEs

Overall, from 2015 – 2022, 397,668 distinct residents filled ≥1 tramadol prescription (Figure 1), with most prescribed an average daily tramadol dose <400mg/day (98.3%) and an average daily MME <50 MME/day (74.3%) (Table 1). Among age groups, more patients were ≥65 years old (32.2%), although high dose (27.1%) and ≥50 MME (28.0%) tramadol use was more common in patients 50 – 64 years old. Overall, females (50.3%) and commercially insured (53.0%) patients were more commonly prescribed tramadol across the dose and MME categories, by largely in-state prescribers (87.2%).

**Figure 1:**
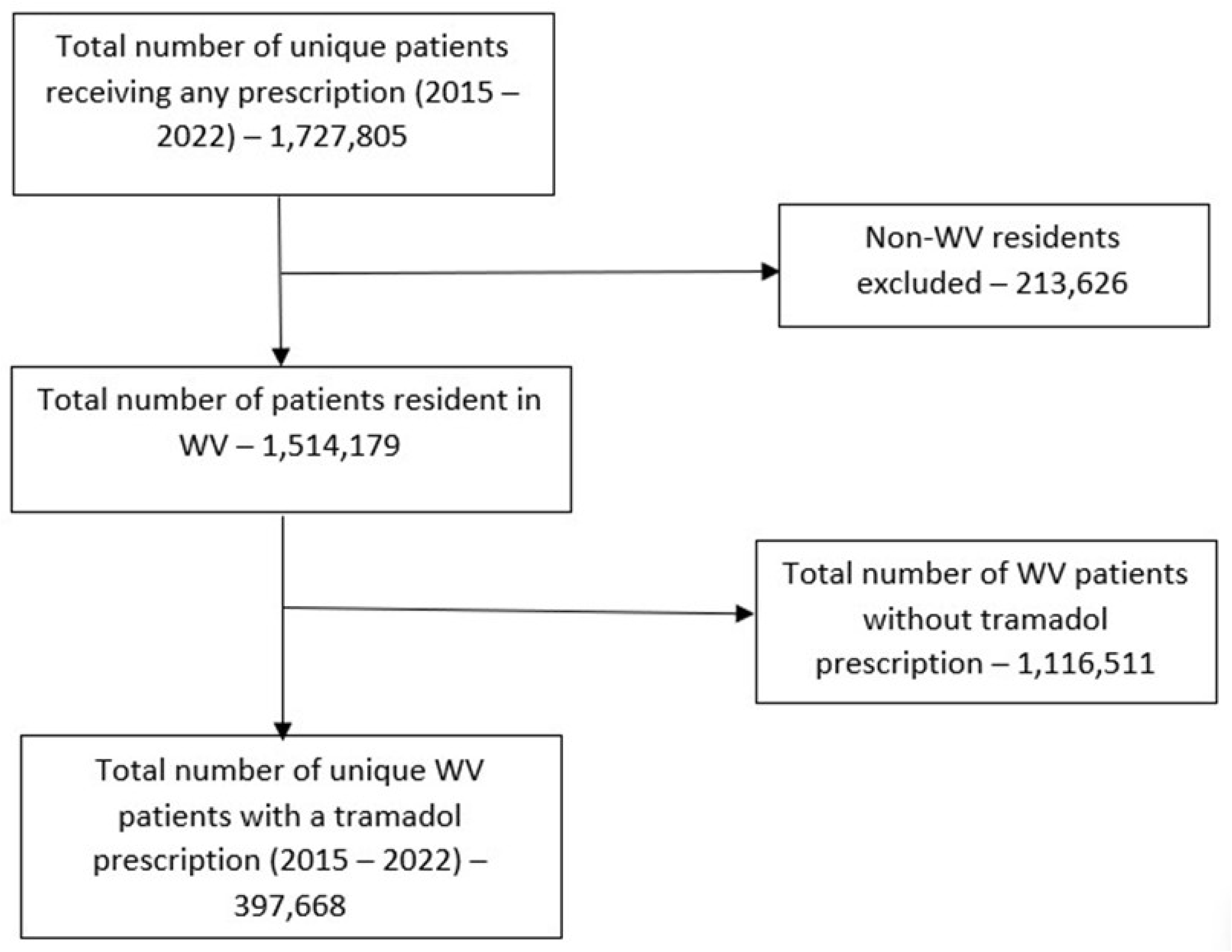
Study Flow Chart

**Table 1:**
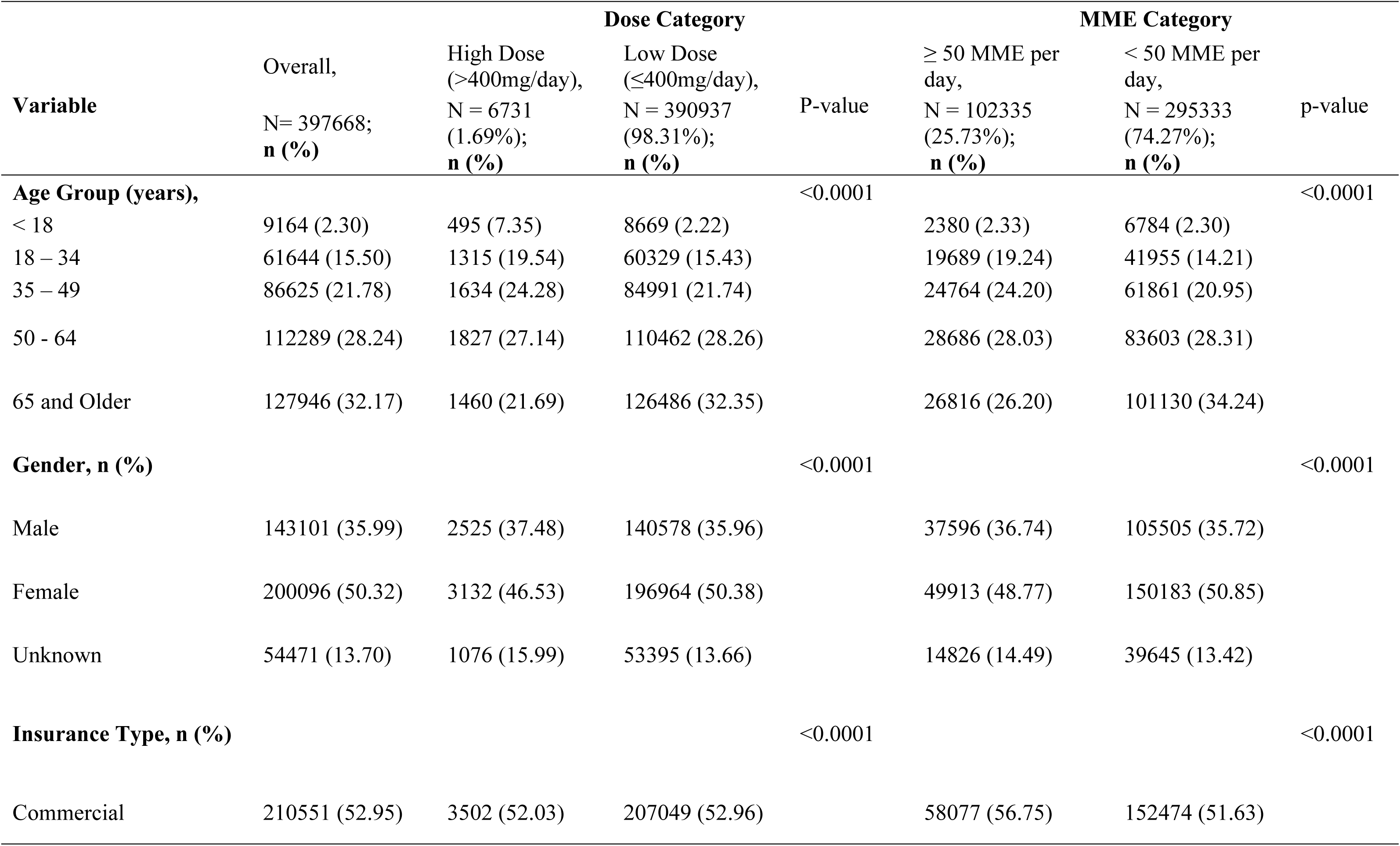
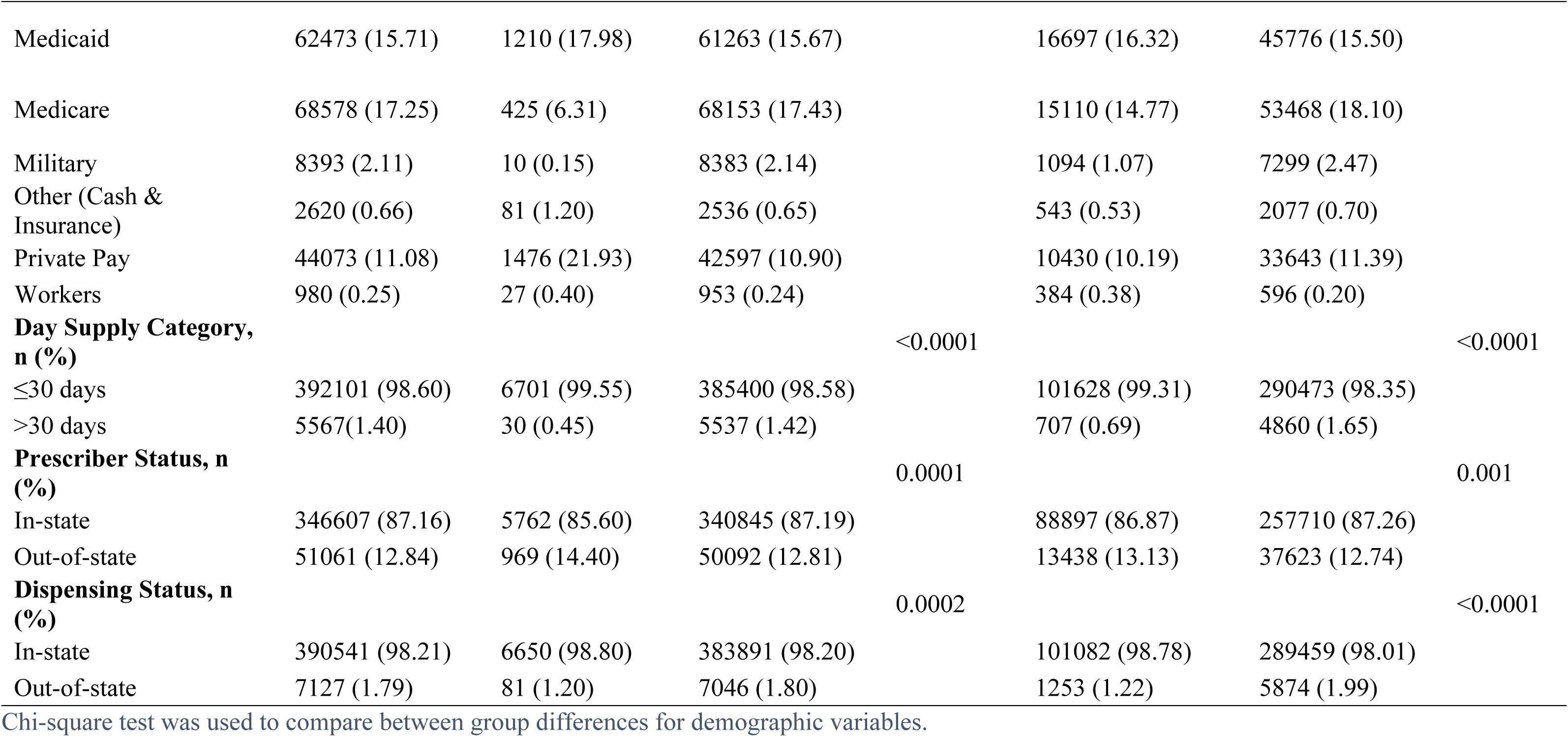
Demographic characteristics of patients with at least one tramadol prescription (2015 – 2022)

### 3.1.2 Patterns of tramadol dispensing

TDR declined gradually by 35% over time (22.1 prescriptions per 100 population in 2015 to 14.4 in 2022), in contrast to ODR which decreased by 57% (111.3 to 48 per 100) within the same period (Figure 2a). A similar trend was observed for tramadol users per 100 population (6.9 – 3.8 per 100 from 2015 to 2022) (Figure 2b).

**Figure 2:**
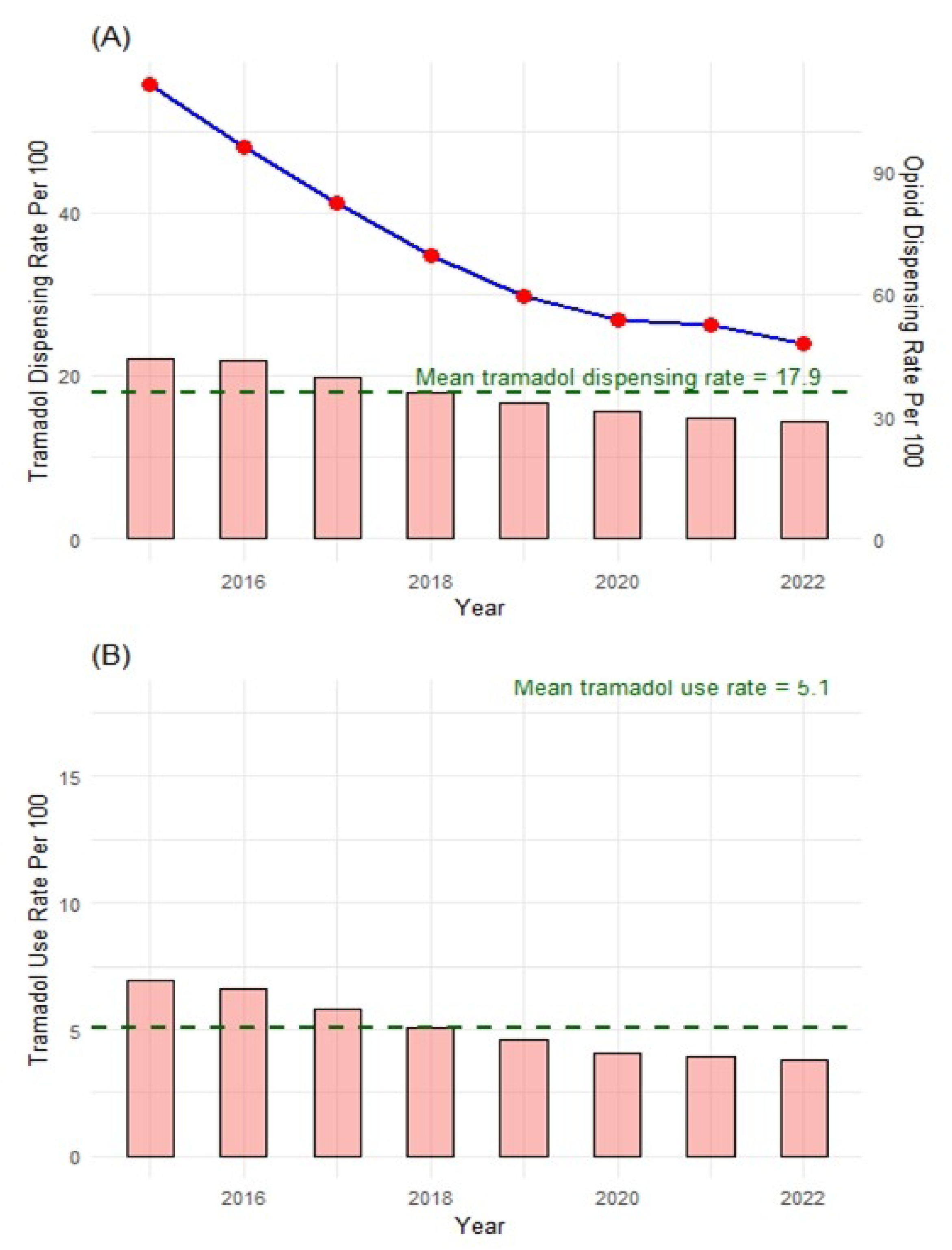
(A) Bar Graph of West Virginia Tramadol Dispensing Rate Per 100 vs Line Plot of West Virginia Opioid Prescription Rate Per 100 Population from 2015 – 2022 (B) Bar Graph of West Virginia Tramadol Use Rate (by person) per 100

County TDR varied widely by county from 2015 to 2022 with Grant County (34.10) having the highest average TDR (Figure 3a). Further, in Figure 3b, mostly southern counties including Mercer (84.9/23.9), McDowell (78.4/28.9), Wyoming (87.3/26.7), and few northern counties (Grant (69.6/34.1), Wetzel (61.7/22.3)) have a high average ODR and a high average TDR. In Pendleton (38.7/30.1), Tucker (32.7/26.2), Lincoln (36.7/24.3), and Hardy (49.8/22.4), a low average ODR and high average TDR were observed (Figure 3b).

**Figure 3a:**
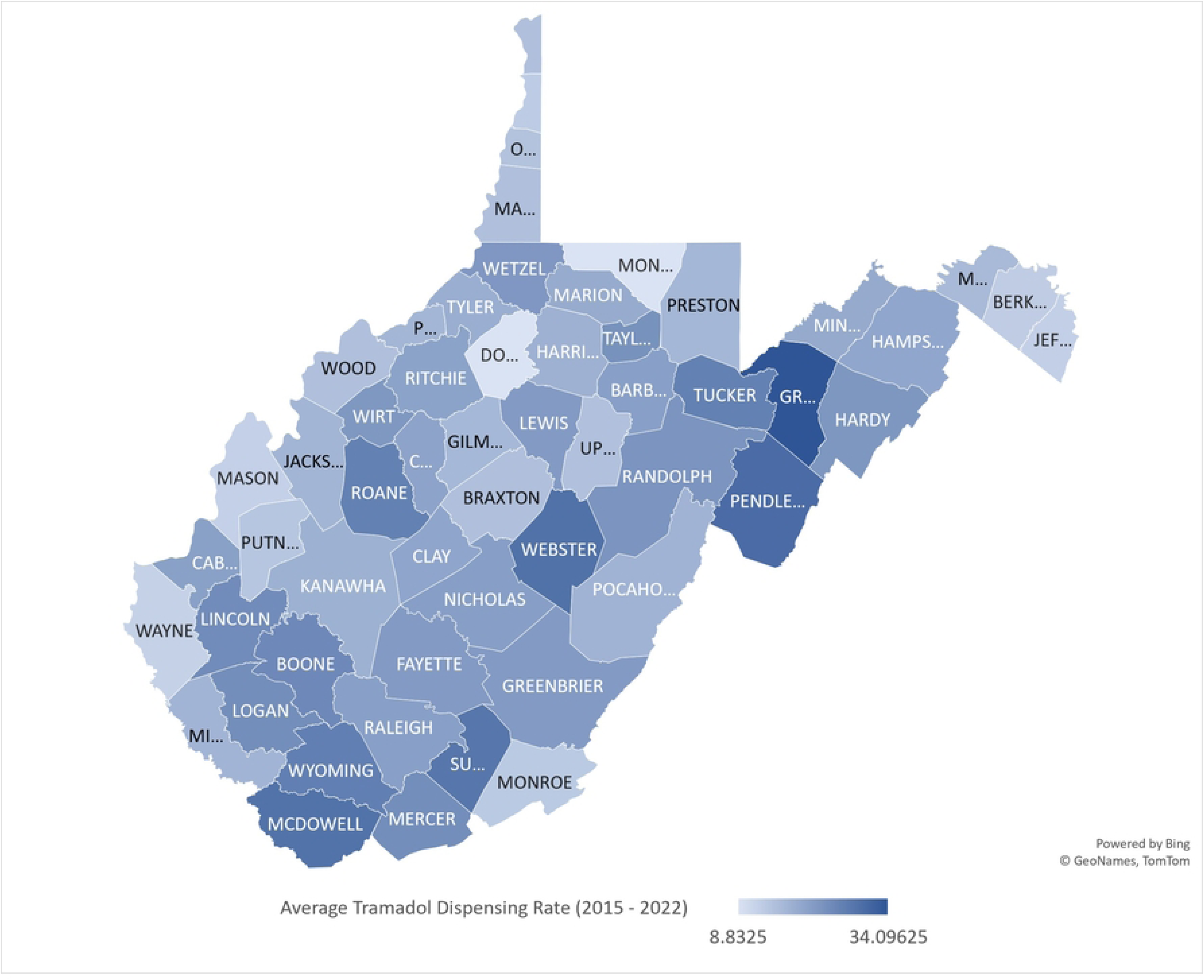
Map of Average Tramadol Dispensing Rate per County (2015 to 2022)

Map lines delineate study areas and do not necessarily depict accepted national boundaries.

**Figure 3b:**
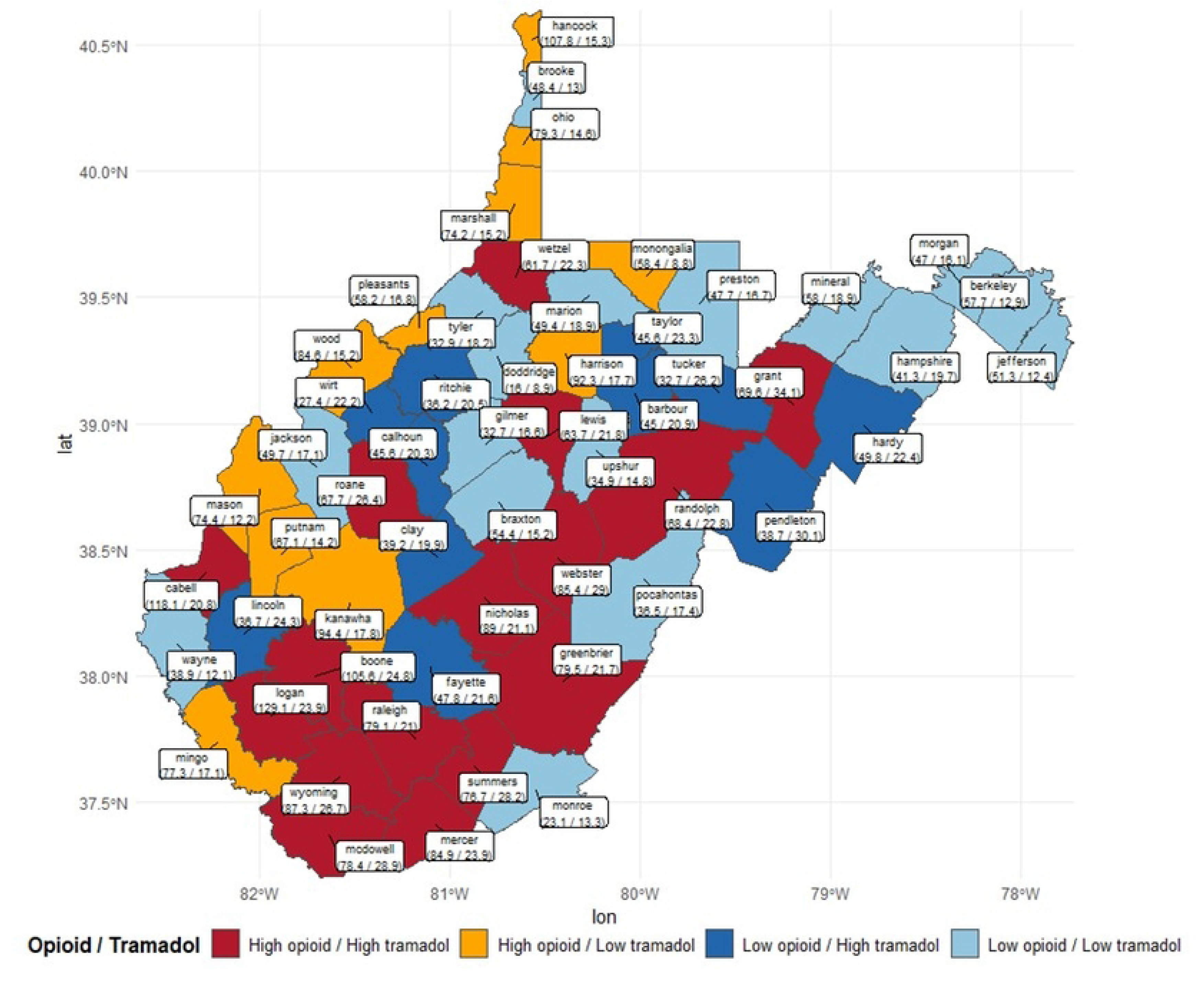
Map of Average Tramadol Dispensing Rate Vs Average Opioid Dispensing Rate by County from 2015 – 2022

Map lines delineate study areas and do not necessarily depict accepted national boundaries.

### 3.1.3 Out of state prescribing and filling of tramadol prescription

Out-of-state prescribers-issued tramadol prescriptions were frequently from Virginia, Ohio, and Maryland (states on a WV border), with fluctuations across years (Supplementary Figure S1a). About 30% of all annual out-of-state tramadol prescriptions originated from Virginia (Supplementary Figure S1a), particularly from Winchester City, Virginia just on the eastern border of WV which had a rising proportion across the years (2015 – 2019) (Supplementary Figure S1b).

Tramadol prescriptions were filled on occasion outside WV in pharmacies located as far away as Arizona and Missouri, as well as in border states of Ohio, Pennsylvania, and Virginia (Supplementary Figure S1c). While approximately 30% of tramadol prescriptions filled in pharmacies outside WV from 2015 – 2018 were in Arizona, specifically Maricopa County (Supplementary Figure S1d), the prescription proportion declined by about 10% in subsequent years (Supplementary Figure S1c). Additional details on out-of-state prescribing and filling of tramadol prescriptions are available in Supplementary Table S1a and S1b.

### 3.1.4 Tramadol co-prescriptions

Yearly, over 30% of patients with a tramadol prescription had concurrent prescriptions for ≥1 controlled substances in the same month, with a slightly increasing or steady proportion across years (Supplementary Figure S2a). More patients (≥10%) were co-prescribed other opioids from 2015 – 2018, later surpassed by gabapentinoids as the most prominent co-prescribed medication from 2019 (Supplementary Figure S2b). Furthermore, when tramadol is co-prescribed with two other drugs, more patients (2 – 5%) had additional prescriptions for benzodiazepines and other opioids (2015 – 2018) or benzodiazepines and gabapentinoids (2019-2022) (Supplementary Figure S2c). Further details on tramadol co-prescription are provided in Supplementary Table S2.

### 3.1.5 Relationship between social determinant of health factors and tramadol dispensing rates across West Virginia

In Table 2, we found positive, significant relationships between higher TDR and the percentage of the county populations with the following variables: poor or fair health (**β = 0.07, p = 0.01),** physical inactivity (**β = 0.10, p = 0.0003**), uninsured (**β = 0.09, p = 0.001)**, living in a rural area (**β = 0.09, p = 0.003**), PCP rate (**β = 0.10, 0.0003**), and high ODR (**β = 0.14, p <0.0001**) above the average/median. Thus, TDR was high in counties with a high proportion of individuals with poor or fair health, physically inactive, or without health insurance. With an existing high OPR or PCP rate, TDR was also elevated.

**Table 2:**
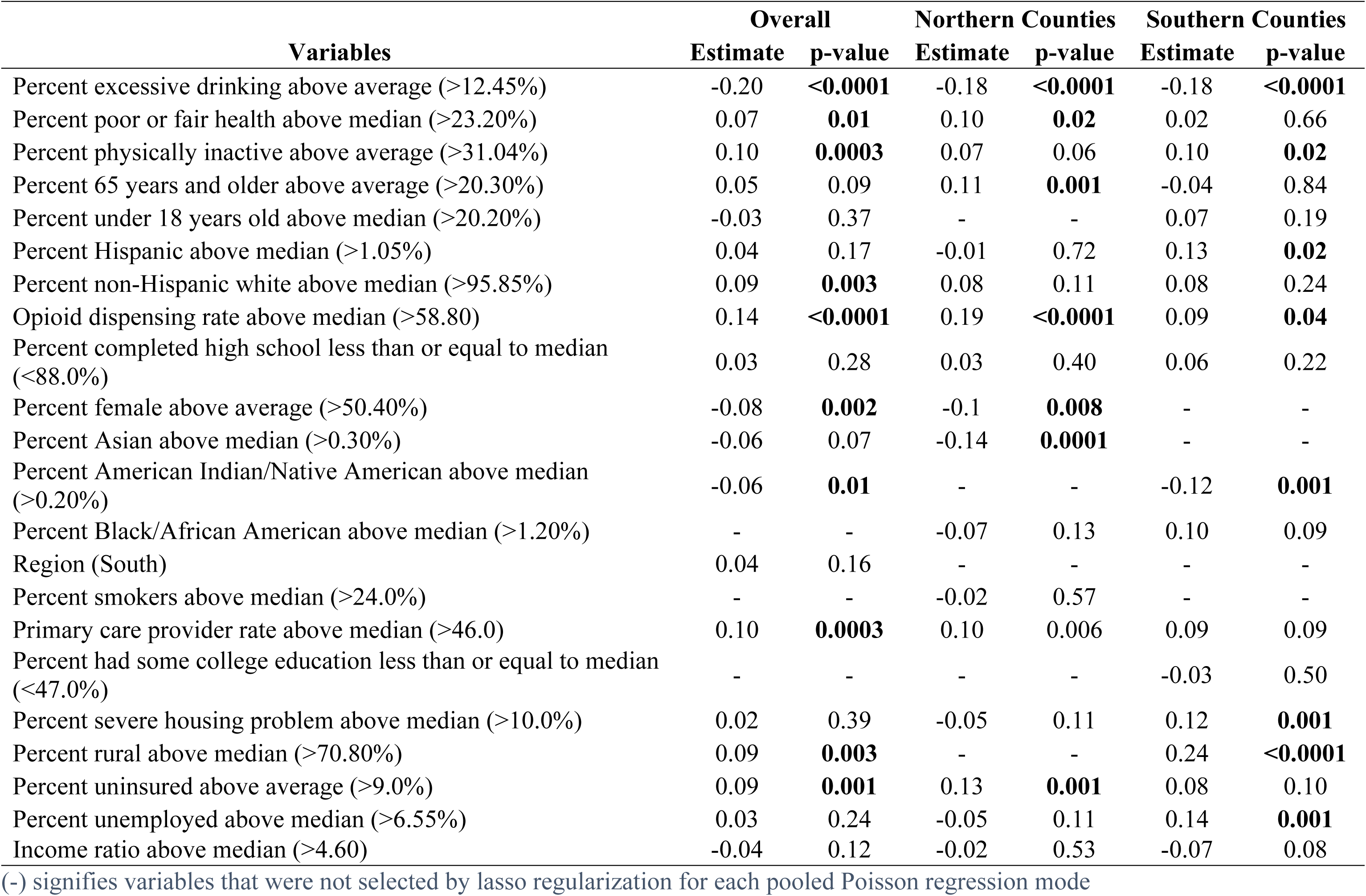
Pooled Poisson Regression of Average Tramadol Dispensing rate per 100 with West Virginia County Social Determinant of Health Factors.

There was a negative statistically significant relationships between TDR and the percentage of the county populations with excessive drinking (**β =-0.20, p < 0.0001),** female (**β =-0.08, p = 0.002),** and American Indian/Native American (**β =-0.06, p = 0.01)** above average/median.

Region-specific analyses had similar findings with a few exceptions. For northern counties, TDR was elevated for counties where the percentage of ≥65 years residents (**β = 0.11, p = 0.001**), uninsured (**β = 0.13, p = 0.001**), and PCP rate (**β = 0.10, p = 0.006**) are above the average/median value. In the southern counties, increased tramadol dispensing rate was observed in counties where percent of population with severe housing problem (**β = 0.12, p = 0.002**), rural (**β = 0.24, p<0.001**), physically inactive (**β = 0.10, p = 0.02**), and unemployed residents (**β = 0.14, p = 0.001**) exceeds the average/median values.

## 4.1 Discussion

Our results did not show an inverse relationship between ODR and TDR, but we discovered county-wise variations in TDR juxtaposed with the respective ODR. TDR gradually declined over time by 35% from 2015 to 2022, but considerably less than the more pronounced 57% decline in ODR. Interestingly, tramadol use per 100 persons also declined across the years. Further, tramadol was more commonly prescribed to older patients at high doses and MMEs. Over 30% of the tramadol users had a concurrent prescription with other opioids, benzodiazepines, and gabapentinoids. Our study found that SDoH factors including proportion of physically inactive, uninsured, unemployed, and frail health, ODR, and PCP rates were associated with increased TDR across the counties.

The decline in TDR post-scheduling is consistent with the trend among commercially insured patients in the U.S. (11) and Rhode Island Medicare Part D enrollees (26). The decline may be a direct or indirect impact of the schedule IV designation and/or implementing PDMP (27–29). In our study, the reduction in TDR was non-commensurate with the decline in ODR by a considerably wide margin (22%). County-wise evaluations revealed variations in prescribing practices, highlighting counties with high ODR/high TDR and low ODR/high TDR, which might be responsible for slowing down state-wide decline in TDR relative to the reduction in state-wide ODR. It is also evident that there was a heterogeneity in prescribing practices within WV. Thus, identifying regions of interest for monitoring and possible prescribing policy reform.

Higher tramadol use rates (e.g., daily tramadol MME intake ≥ 50 MME) within older patients can be explained by a higher prevalence of arthritis (32.8%, age adjusted) in WV in 2015, highest in the Appalachian regions (30). Also, 20.5% WV population had multiple chronic conditions, which may also contribute to chronic pain (31). Additionally, higher tramadol utilization among females aligns with other studies, though the rationale behind it is unclear (6,11,32,33). Additional studies are needed to explore driving factors for higher tramadol use in females while accounting for possible sex differences in pain perception and other prescribing patterns.

About 13% out of state tramadol prescriptions were probably through mail in order, accounting for about 23,000 – 50,000 tramadol prescriptions per year. This may be further assessed to ensure drug diversion does not occur. Moreover, among concurrent prescriptions with tramadol, the rise in gabapentinoid prescriptions can be attributed to the designation of gabapentin as a schedule V drug in 2018 (34). The proportions of co-users reported by other studies are comparable to our finding of 3 - 30% (11,33). The co-prescription of opioids and benzodiazepines is not recommended except in patients without a suitable alternative due to increased risks of overdose and adverse effects like falls, respiratory depression, and death (35,36). There have been reports of tramadol-involved deaths in the presence of either therapeutic or lethal benzodiazepine levels (37). Tramadol co-use with gabapentin was also implicated in a case of serotonin syndrome which presented with confusion, agitation, aggression, and tachycardia (38).

The heterogeneity in TDR varied by WV county, with Grant, Wyoming, and McDowell counties exceeding the dispensing rate for the entire state. According to BRFSS reports (2015 – 2019), Wyoming and McDowell counties have higher arthritis prevalence compared to the state’s prevalence (13), which could imply a high pain burden in both counties and might be linked to their high tramadol dispensing rates. Counties with high opioid/high tramadol and low opioid/high tramadol dispensing rates including Summer, Grant, and Pendleton, have higher proportion of adults 65 years and older (39). High tramadol dispensing rate in Wyoming and McDowell counties is further complicating the polysubstance epidemic since both counties rank high in vulnerability to all-drug overdose mortality in the state (40). Although Grant county has a low all-drug overdose death vulnerability (40), it has had a rate of opioid analgesics per 1000 population, proportion of patients receiving >90 MME of opioids, and overlapping opioid and benzodiazepine prescriptions comparable to the entire state’s rates and proportions from 2014 to 2022 (41). Thus, these counties need to be closely monitored for tramadol utilization to circumvent probable drug overdose related problems.

Our study revealed some important association between SDoH and high TDR in certain counties. These relationships were unexplored in previous national and state-level studies on tramadol utilization (11,26).

However, certain SDoH factors have often been implicated in high utilization of prescription opioids within different subsets of patients, raising significant issues and potentially modifiable factors for health optimization. These studies reported an association between prescription opioid utilization and low physical activity, unemployment, poor health status, primary care physician shortage area status, and less than high school education, which concur with our findings (42–46). The proportion of Black residents within a predominantly White residents region had a positive association with opioid utilization, (42) which corresponds with our findings of higher tramadol utilization in southern WV counties with a high proportion of Black/African American residents. The region-based analysis identified severe housing problem, rurality, and unemployment as significant predictors of high tramadol use prominent in southern WV unlike northern WV. Thus location-targeted monitoring, policies, and interventions are necessary for tramadol utilization control.

Our study has many implications in monitoring and preventing overdose deaths related to tramadol within WV. Addressing differences in SDoH holds promise for a reduced disease burden, better health, reduced health resource need and utilization, and lesser individual and payer-level costs. Quantifying these associations and identifying regions of the state with high TDR offers insight not only on locations in need of closer monitoring but also factors for targeted intervention for health improvement and reduced drug utilization. These regions would also benefit from more stringent PDMP practices for continuous monitoring of tramadol utilization and preventing probable misuse. Highlighting demographic variabilities such as greater female tramadol use identifies areas in need of further study. Patterns of high-dose tramadol use and co-prescription with other controlled substances indicates the need for awareness of the possibility of adverse events or drug misuse. Consequently, safe tramadol co-prescription practices would be required of prescribers to ensure safety and patients/caregivers should be duly notified and educated on safety concerns and provided with access to helpful resource when tramadol is co-prescribed with prescription medicines capable of drug-drug interaction.

The strengths of this study include the detailed analysis of tramadol utilization in WV at both state and county levels. Unlike prior studies focused on either commercially insured or Medicare populations, (11,26) this study included tramadol users irrespective of age, sex, and insurance type. By linking county-level tramadol utilization with the county health ranking and roadmap data, we could quantify the relationship between the TDR and SDoH. Inclusion of medications co-prescribed with tramadol showed prescription patterns changes over time. This study also had some limitations. First, we did not explore changes in TDR within the demographic groups, e.g., age, sex, and insurance types over the years. Thus, it was not evident how tramadol utilization could have changed within these groups. Potentially important demographic traits such as race/ethnicity, income status, level of education, and marital status which might impact tramadol utilization were unavailable in the dataset and not described. There was no information on the indication for the tramadol prescriptions or comorbidities in these patients, so the disease burden within the study population and need for a tramadol prescription could not be determined. Finally, information on prescriber specialty was not available in the dataset, so, we were unable to link specialties with the volume of tramadol prescriptions generated.

## 4.2 Conclusion

Following its schedule IV designation, tramadol utilization declined in WV, yet this overall drop masks striking variations in usage pattern across counties, sexes, age groups, and insurance types. County-wise differences in tramadol utilization were found associated with a high proportion of uninsured, unemployed, physically inactive, and frail health residents and a high opioid dispensing rate and patient to provider ratio. Addressing these factors might positively improve health, reduce disease burden, decrease drug and health resource utilization, as well as costs for individuals, third-party payers, and the state.

Future studies should explore the reasons for greater use of tramadol in females and variations in tramadol utilization across demographic strata, as well as the long-term impact, if any, of concomitant tramadol use with other controlled substances on drug dependence, drug misuse, and adverse effects.

## Data Availability

Data cannot be shared publicly because of its confidential nature and restrictions.

## Abbreviations

CSA: Controlled Substances Act
DEA: Drug Enforcement Administration
ED: Emergency Department
NSUDH: National Survey of Drug Use and Health
NFLIS: National Forensic Laboratory Information System
WV: West Virginia
BRFSS: Behavioral Risk Factor Surveillance System
WVCSMP: West Virginia Controlled Substances Monitoring Program
NDC: National Drug Code
IRB: Institutional Review Board CDC – Centers for Disease Control
MME: Morphine Milligram Equivalent TDR – Tramadol Dispensing Rate
ODR: Opioid Dispensing Rate PCP – Primary Care Provider

## Acknowledgements

N/A

## Funding

The project was supported by the National Institute of General Medical Sciences [Grant number - U54GM104942].

## Declaration of interest statement

The authors have no conflicting financial interests or personal relationships that could have affected the work in this publication.

## Data availability statement

The participants of this study did not give written consent for their data to be shared publicly, so due to the sensitive nature of the research supporting data is not available.

## Supplementary Information

Figure S1:

(A) Top 3 States with the Highest Proportion of Tramadol Prescriptions from Prescribers Outside West Virginia

(B) Top 3 Counties with the Highest Proportion of Tramadol Prescriptions from Prescribers Outside West Virginia

(C) Top 3 States with the Highest Proportion of Tramadol Prescriptions Filled in Pharmacies Outside West Virginia

(D) Top 3 Counties with the Highest Proportion of Tramadol Prescriptions Filled in Pharmacies Outside West Virginia

The proportions were calculated for Figure 4A & 4B using the total prescriptions from each state or county as a percentage of the total number of tramadol prescriptions written by an out-of-state prescriber in each year (2015 N = 50062, 2016 N = 45576, 2017 N = 38719, 2018 N =30371, 2019 N = 25219, 2020 N = 23926, 2021 N = 24099, 2022 N = 23852). The proportions were calculated for Figure 4C & 4D using the total prescriptions from each state or county as a percentage of the total number of tramadol prescriptions filled in an out-of-state pharmacy in each year *(2015 N = 8888, 2016 N = 8775, 2017 N = 8898, 2018 N = 8102, 2019 N = 9218, 2020 N = 11429, 2021 N = 12691, 2022 N = 13310*)

Table S1a: Percentage of Tramadol Prescriptions Issued by an Out-of-State Prescriber

Table S1b: Percentage of Tramadol Prescriptions Filled in an Out-of-State Pharmacy

Figure S2: Tramadol Co-Prescription by Number & Type Per Year The percentage was calculated using the number of patients who had multiple prescriptions including tramadol in the same month of a particular year and the total number of patients prescribed tramadol for that year (2015 N = 127089, 2016 N = 120078, 2017 N = 105148, 2018 N = 91044, 2019 N = 81392, 2020 N = 71897, 2021 N = 69314, 2022 N = 66953)

Table S2: Percentage of Patients with Tramadol Co-Prescription by Number and Type of Prescription Medicines

## Notes

### Competing Interest Statement

The authors have declared no competing interest.

### Funding Statement

Yes

### Author Declarations

Institutional Review Board West Virginia University

## References

1. Dhesi M, Maldonado KA, Patel P, Maani CV. Tramadol. In: StatPearls [Internet]. Treasure Island (FL): StatPearls Publishing; 2024 [cited 2024 June 25]. Available from: http://www.ncbi.nlm.nih.gov/books/NBK537060/

2. Dunn KE, Bergeria CL, Huhn AS, Strain EC. A Systematic Review of Laboratory Evidence for the Abuse Potential of Tramadol in Humans. Front Psychiatry. 2019 Sept 26;10:704.

3. Federal Register [Internet]. 2013 [cited 2024 Sept 24]. Schedules of Controlled Substances: Placement of Tramadol Into Schedule IV. Available from: https://www.federalregister.gov/documents/2013/11/04/2013-25933/schedules-of-controlled-substances-placement-of-tramadol-into-schedule-iv

4. Parisi T. Addiction Center. 2024 [cited 2024 June 24]. Tramadol Addiction And Abuse. Available from: https://www.addictioncenter.com/opiates/tramadol/

5. Federal Register [Internet]. 2014 [cited 2024 June 25]. Schedules of Controlled Substances: Placement of Tramadol Into Schedule IV. Available from: https://www.federalregister.gov/documents/2014/07/02/2014-15548/schedules-of-controlled-substances-placement-of-tramadol-into-schedule-iv

6. Mullins PM, Mazer-Amirshahi M, Pourmand A, Perrone J, Nelson LS, Pines JM. Tramadol Use in United States Emergency Departments 2007-2018. J Emerg Med. 2022 May;62(5):668–74.

7. 7. Drug Enforcement Administration Diversion Control Division Drug & Chemical Evaluation Section. Tramadol (Trade Names: Ultram®, Ultracet®) [Internet]. Drug Enforcement Administration. 2025 [cited 2025 July 23]. Available from: https://www.deadiversion.usdoj.gov/drug_chem_info/tramadol.pdf

8. Key Substance Use and Mental Health Indicators in the United States: Results from the 2022 National Survey on Drug Use and Health. 2022;

9. NFLIS HOME [Internet]. 2024 [cited 2024 July 8]. Available from: https://www.nflis.deadiversion.usdoj.gov:8443/

10. NFLIS Publications [Internet]. 2024 [cited 2024 July 8]. Available from: https://www.nflis.deadiversion.usdoj.gov:8443/publicationsRedesign.xhtml

11. Basham CA, Edrees H, Huybrechts KF, Hwang CS, Bateman BT, Bykov K. Tramadol use in U.S. adults with commercial health insurance, 2005-2021. American Journal of Preventive Medicine [Internet]. 2024 June 12 [cited 2024 June 28]; Available from: https://www.sciencedirect.com/science/article/pii/S0749379724001995

12. WV Medicaid Age Limits for Tramadol-Containing Agents [Internet]. 2025 [cited 2025 July 22]. Available from: https://dhhr.wv.gov/bms/BMS%20Pharmacy/Pages/WV-Medicaid-Age-Limits-for-Tramadol-Containing-Agents.aspx

13. Behavioral Risk Factor Surveillance System [Internet]. 2019 [cited 2024 Sept 5]. Available from: https://dhhr.wv.gov/HSC/SS/BRFSS/Pages/BRFSS.aspx

14. Disability Statistics [Internet]. 2022 [cited 2025 Feb 4]. Available from: https://www.disabilitystatistics.org/

15. Explore Arthritis in West Virginia - AHR [Internet]. 2024 [cited 2025 Feb 4]. Available from: https://www.americashealthrankings.org/explore/measures/Arthritis/Arthritis_difmob/WV

16. West Virginia State Profile [Internet]. 2024 [cited 2025 Feb 4]. Available from: https://www.umt.edu/rural-disability-research/focus-areas/disability_maps/state-maps/west-virginia.php

17. West Virginia Office of Epidemiology and Prevention Services [Internet]. 2022 [cited 2025 Feb 4]. Available from: https://oeps.wv.gov/Pages/Search.aspx?q=cancer%20burden%20report%202022

18. West Virginia Board of Pharmacy—Controlled Substance Monitoring Program [Internet]. West Virginia Department of Health and Human Resources; 2024 [cited 2025 July 16]. 2023 Controlled Substance Prescribing in West Virginia: County Level Patient Data. Available from: https://dhhr.wv.gov/vip/county-reports/Documents/2023%20CSMP%20Reports/2023%20CSMP%20Surveillance%20Maps%20Report_Final.pdf

19. CDC. Overdose Prevention. 2024 [cited 2024 July 8]. Opioid Dispensing Rate Maps. Available from: https://www.cdc.gov/overdose-prevention/data-research/facts-stats/opioid-dispensing-rate-maps.html

20. Bureau UC. Census.gov. 2025 [cited 2024 July 8]. Census.gov. Available from: https://www.census.gov/en.html

21. Health Data - County Health Rankings & Roadmaps [Internet]. 2024 [cited 2024 Sept 7]. Available from: https://www.countyhealthrankings.org/health-data

22. Dowell D. CDC Clinical Practice Guideline for Prescribing Opioids for Pain — United States, 2022. MMWR Recomm Rep [Internet]. 2022 [cited 2024 June 22];71. Available from: https://www.cdc.gov/mmwr/volumes/71/rr/rr7103a1.htm

23. Buuren SV, Groothuis-Oudshoorn K. mice: Multivariate Imputation by Chained Equations in R. J Stat Soft [Internet]. 2011 [cited 2025 July 22];45(3). Available from: http://www.jstatsoft.org/v45/i03/

24. Judicial Districts by County, Southern District of West Virginia, United States District Court [Internet]. 2025 [cited 2024 Oct 21]. Available from: https://www.wvsd.uscourts.gov/court-info/judicial-districts-county

25. R: The R Project for Statistical Computing [Internet]. 2025 [cited 2025 July 16]. Available from: https://www.r-project.org/

26. Borrelli EP. Assessing the Trends of Tramadol Utilization in the Medicare Part D Population in Rhode Island. R I Med J (2013). 2024 May 2;107(5):33–7.

27. D’Souza RS, Lang M, Eldrige JS. Prescription Drug Monitoring Program. In: StatPearls [Internet]. Treasure Island (FL): StatPearls Publishing; 2025 [cited 2025 Apr 3]. Available from: http://www.ncbi.nlm.nih.gov/books/NBK532299/

28. Tran S, Lavitas P, Stevens K, Greenwood BC, Clements K, Alper CJ, et al. The Effect of a Federal Controlled Substance Act Schedule Change on Hydrocodone Combination Products Claims in a Medicaid Population. JMCP. 2017 May;23(5):532–9.

29. Gupta S, Nguyen T, Freeman PR, Simon K. Competitive effects of federal and state opioid restrictions: Evidence from the controlled substance laws. Journal of Health Economics. 2023 Sept 1;91:102772.

30. Schmajuk G, Trupin L, Yelin E, Blanc PD. Prevalence of Arthritis and Rheumatoid Arthritis in Coal Mining Counties of the U.S. Arthritis Care Res (Hoboken). 2019 Sept;71(9):1209–15.

31. America’s Health Rankings [Internet]. 2025 [cited 2025 Sept 10]. Explore Multiple Chronic Conditions in West Virginia | AHR. Available from: https://www.americashealthrankings.org/explore/measures/CHC/WV

32. Bigal LM, Bibeau K, Dunbar S. Tramadol Prescription over a 4-Year Period in the USA. Curr Pain Headache Rep. 2019 Aug 6;23(10):76.

33. Eric P. Borrelli P, Jeffrey Bratberg P, Mary L. Greaney P, Stephen J. Kogut P. Tramadol utilization among patients with higher risk of adverse drug events: A claims analysis of commercially insured and Medicare Advantage members. Journal of Opioid Management. 2023 Apr 19;19(3):257–71.

34. West Virginia Code Section 60A-2-212 [Internet]. West Virginia Code. 2025 [cited 2025 July 1]. Available from: https://code.wvlegislature.gov/60A-2-212/

35. American Geriatrics Society 2023 updated AGS Beers Criteria® for potentially inappropriate medication use in older adults. 2023 [cited 2025 Apr 4]; Available from: https://agsjournals.onlinelibrary.wiley.com/doi/10.1111/jgs.18372

36. U.S. Food & Drug Administration. FDA Drug Safety Communication: FDA warns about serious risks and death when combining opioid pain or cough medicines with benzodiazepines; requires its strongest warning. FDA [Internet]. 2024 Aug 26 [cited 2025 Apr 4]; Available from: https://www.fda.gov/drugs/drug-safety-and-availability/fda-drug-safety-communication-fda-warns-about-serious-risks-and-death-when-combining-opioid-pain-or

37. Clarot F, Goullé JP, Vaz E, Proust B. Fatal overdoses of tramadol: is benzodiazepine a risk factor of lethality? Forensic Sci Int. 2003 June 24;134(1):57–61.

38. Ekşi MŞ, Turgut VU, Özcan-Ekşi EE, Güngör A, Tükel Turgut FN, Pamir MN. Serotonin Syndrome Following Tramadol and Gabapentin Use After Spine Surgery. World Neurosurgery. 2019 June 1;126:261–3.

39. National Institute of Minority Health and Health Disparities. Population Table for West Virginia Counties | HDPulse Data Portal [Internet]. 2025 [cited 2025 Aug 15]. Available from: https://hdpulse.nimhd.nih.gov/data-portal/social/table?socialtopic=070&socialtopic_options=social_6&demo=00028&demo_options=pop_12&race=00&race_options=raceall_1&sex=0&sex_options=sex_3&age=999&age_options=ageNA_1&statefips=54&statefips_options=area_states

40. County-level vulnerability to overdose deaths in West Virginia [Internet]. Office of Epidemiology & Prevention Services; 2019 [cited 2025 June 24]. Available from: https://oeps.wv.gov/hcv/documents/data/WV_OD_Vulnerability_Assessment.pdf

41. West Virginia Board of Pharmacy Prescription Opioid Indicators Report Grant County - 2022 [Internet]. West Virginia Violence and Injury Prevention Program (WVVIPP); 2022 [cited 2025 June 24]. Available from: https://dhhr.wv.gov/vip/county-reports/CountyReports/County%20Reports%202022/Grant%2022.pdf

42. Moriya AS, Xu L. The complex relationships among race/ethnicity, social determinants, and opioid utilization. Health Serv Res. 2021 Apr;56(2):310–22.

43. Pratap P, Dickson A, Love M, Zanoni J, Donato C, Flynn MA, et al. Public Health Impacts of Underemployment and Unemployment in the United States: Exploring Perceptions, Gaps and Opportunities. Int J Environ Res Public Health. 2021 Sept 23;18(19):10021.

44. Wague A, O’Donnell JM, Rangwalla K, El Naga AN, Gendelberg D, Berven S. Impact of social determinants of health on perioperative opioid utilization in patients with lumbar degeneration. North American Spine Society Journal (NASSJ). 2023 June 1;14:100221.

45. Zajacova A, Grol-Prokopczyk H, Limani M, Schwarz C, Gilron I. Prevalence and correlates of prescription opioid use among US adults, 2019–2020. PLOS ONE. 2023 Mar 2;18(3):e0282536.

46. Zajacova A, Lawrence EM. The relationship between education and health: reducing disparities through a contextual approach. Annu Rev Public Health. 2018 Apr 1;39:273–89.

